# Trends in Suicide Mortality by Method among US Individuals aged 10-24 Years from 1999 to 2024

**DOI:** 10.64898/2026.06.12.26355540

**Authors:** Anne Bischops, Marie-Laure Charpignon, Kenneth D. Mandl, Maimuna Majumder

**Author notes:** These authors contributed equally and share senior authorship (Kenneth D. Mandl, Maimuna Majumder). Corresponding author: Dr. Anne C. Bischops, Computational Health Informatics Program, Boston Children’s Hospital, 300 Longwood Avenue, Boston, MA 02115, USA.

## Abstract

**Background:** Suicide is the second leading cause of death in US adolescents aged 10-24. Method use strongly influences lethality and design of prevention strategies, but recent trends remain unclear. We therefore aimed to investigate trends in suicide mortality rates by method, age group, and sex.

**Methods:** This cross-sectional study used suicide mortality data from the National Center for Health Statistics for a quarter-century period, between 1999 and 2024. All individuals aged 10-24 years at the time of death, with suicide as the underlying cause, were included. We estimated suicide mortality rates (i.e., the number of suicide deaths per 100,000 people) and annual percent change by method (firearm, asphyxiation, poisoning, other), age group (10-14, 15-19, 20-24), and sex. Changing trend time points were determined using Joinpoint regression models

**Results:** From 1999 to 2024, 159,241 suicide deaths occurred among individuals aged 10-24. While suicide rates declined across all age groups between 2017 and 2024, the male-to-female gap narrowed by 18.9%. Among 10-14-year-olds, declining rates among males masked a consistent increase in female suicide rates since 2011. Although asphyxiation-related suicides decreased across all groups since 2018, firearm suicide rates increased for females in the 10-14 and 20-24 age groups. Albeit not as common as firearms or asphyxiation, poisoning suicide rates increased in the 15-19 and 20-24 age groups. Since 1999, suicide rates by other less common methods (e.g., jumping) showed significant increases, for both sexes, especially among individuals aged 20-24. Suicide rates were consistently highest in the 20-24 age group across all study years.

**Conclusion:** The decrease in suicide mortality rates among individuals aged 10-24 was largely driven by declines in males and reductions in asphyxiation-related suicides. However, increasing female suicide rates in the 10-14 age group, as well as increasing rates of death by less common means, warrant close attention. While suicide prevention efforts like structural interventions and means restriction have shown effectiveness among male adolescents, priority should now be given to adapting these approaches for female adolescents, particularly those aged 10-14.

## Introduction

The Lancet Commission on adolescent health projects that, by 2030, mental health disorders and suicides will account for the loss of 42 million healthy life-years among adolescents.^1^ Since 2010, suicide has been the second leading cause of death after accidents among individuals aged 10-24 in the United States (US)—inclusive of a 46% increase between 1999 and 2019.^2–5^

The COVID-19 pandemic profoundly exacerbated adolescent mental health. In 2020, the absolute count of adolescent suicide deaths increased in 14 US states, and in 2021, one in three adolescents reported poor mental health.^6,7^ In response, the American Academy of Pediatrics declared a national emergency in adolescent mental health.^8^ Nevertheless, in 2023, 29% of adolescents aged 14-18 years continued to experience poor mental health, and 10% had attempted suicide.^9^ These alarming figures suggest that protective measures have been insufficient and underscore the urgent need for prevention strategies targeted towards high-risk subgroups.

Identifying these subgroups requires understanding who is most at risk and how risk has shifted over time. Historically, young adult males have had higher suicide mortality rates than their female counterparts.^3,4^ However, the 2010s were marked by a rapid increase in female suicides.^3,4^ As of 2021, the number of suicide attempts among girls aged 12-17 was 51% higher than in 2019, suggesting a shift in previously identified sex-based patterns.^10^ Moreover, suicide risk is affected by access to lethal means.^4^ The choice of suicide method strongly influences both the likelihood of death and the types of prevention strategies that can be effective, making it essential to understand which methods are most commonly used and by whom.^4^ Among 10-19-year-olds, firearm and asphyxiation were the leading methods of suicide until 2020. Firearm-related suicides rose steadily from 2007 to 2020, while asphyxiation-related suicides—despite remaining a leading method throughout this period—began declining after 2018.^4^ Meanwhile, for the 20-24 age group, suicide rates increased until 2017, yet little is known about recent trends in method choice.^4^

Whether the pandemic further disrupted these pre-2020 patterns and whether emerging trends vary by method, age, and sex remains largely underexplored. Updated, nationally representative data on both temporal changes and demographic differences in method choice are critical to informing effective suicide prevention strategies. Therefore, we sought to examine patterns of suicide mortality rates among individuals aged 10-24 from 1999 to 2024—the most extensive timeframe examined in the literature to date—with a particular focus on how method use varied by age group and sex over time.

## Methods

This cross-sectional study examined national-level suicide mortality rates from 1999 to 2024 by method, using mortality data from the National Center for Health Statistics (NCHS) publicly available via the Centers for Disease Control and Prevention’s Wide-ranging Online Data for Epidemiologic Research platform (CDC WONDER).^11^ Our analysis focused on suicide as the underlying cause of death. We examined ages 10-24, in line with the Lancet Commission’s updated definition of adolescence.^1^

Four suicide method categories were considered: firearms, poisoning, asphyxiation (including hanging), and other (including jumping and self-harm by a sharp object), based on the International Statistical Classification of Diseases and Related Health Problems, Tenth Revision (ICD-10) codes. For each method, both crude and age-standardized suicide mortality rates were estimated (expressed in terms of events per 100,000 people), overall and by subgroup. Specifically, crude death rates among individuals aged 10-24 were age-standardized to the US population in 2000, using the direct method as well as a stratification into three age groups (10-14, 15-19, 20-24). Further, we estimated suicide mortality rates by age group (10-14, 15-19, 20-24) and by sex. In addition, the share of suicide deaths, out of all adolescent deaths (in %), and the share of suicide deaths by method were calculated. Of note, death counts <10 are suppressed by the NCHS to protect personal privacy and prevent identification. We chose suicide method and age group categories accordingly to ensure robust and interpretable results as well as comparability with previous work.

Building on prior research by Ormiston, Miron, and Hedegaard,^3–5^ changing trend time points were determined using joinpoint regression models. Annual percent change (APC) estimates were derived from these models, with a significance threshold of p<0.05 (2-tailed t-test). The type of joinpoint model and the number of joinpoints were selected based on the minimum value of the weighted Bayesian Information Criterion (BIC). Average APCs (AAPCs) were further calculated for all subgroups for the entire study period (1999-2024) and for the most recent years (2020-2024). The statistical software R (version 4.5) and the National Cancer Institute’s Joinpoint Regression program (version 5.4) were used.^12,13^

Data analysis was performed from January 15 to February 15, 2026. Study findings were reported in adherence to the Strengthening the Reporting of Observational Studies in Epidemiology (STROBE) Guidelines.^14^

## Results

### Trends in suicide mortality among individuals aged 10-24 years

Between 1999 and 2024, 159,241 suicide deaths occurred among individuals aged 10-24 in the US. In 2024, suicides accounted for 19.1% of adolescent deaths (Figure S1), and the crude suicide mortality rate (CR) was 9.7 per 100,000 adolescents (95% CI: 9.5 to 10.9). Since 2017, suicide mortality rates decreased among all age groups. For the 10-14 age group, suicide rates decreased on average by 3.0 (CI: −7.2 to −0.1; p=0.047) over the 2020-2024 period. For the 15-19 age group, the AAPC over the 2020-2024 period was estimated at −2.7 (CI: −4.2 to −1.5; p=0.002). Similarly, for the 20-24 age group, the AAPC was estimated at −2.0 (CI: −3.6 to −0.9; p=0.014) (Figure 2).

**Figure 2:**
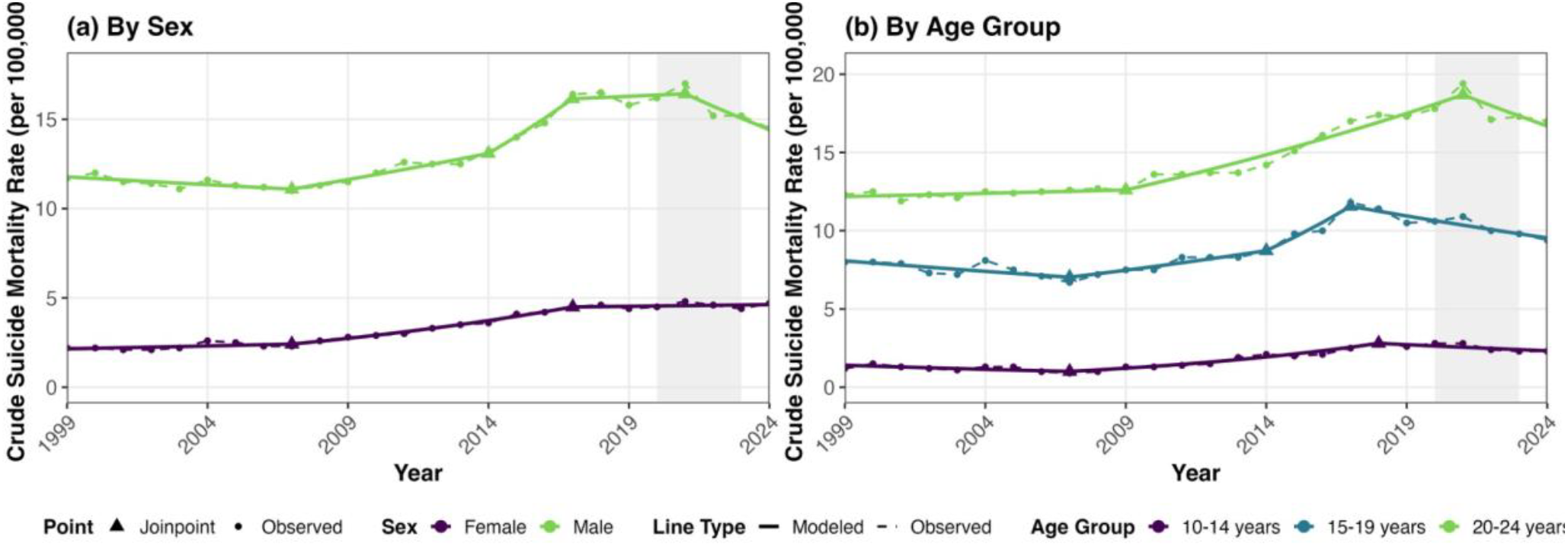
Joinpoint regression of crude suicide mortality rates by (a) sex and (b) age group for individuals aged 10-24. The grey shading denotes the COVID-19 emergency period (2020-2023) for the US.

From 1999 to 2017, suicide mortality rates were four times higher among males (CR: 12.3 (CI: 11.9-12.7)) than among females (CR: 2.9 (CI: 2.7-3.1)). This gap between sexes decreased by 18.9% between 2017 and 2024 (CR males: 15.8 (CI: 15.3-16.2), CR females: 4.6 (4.3-4.8)). While suicide rates decreased on average by 3.1 (CI: −4.2 to −2.1, p <0.001) among males over the 2020-2024 period, no change occurred among females, with an AAPC of 0.4 (CI: −1.4 to 1.7, p=0.554) (Figure 2).

Importantly, suicide mortality rates among females aged 10-14 substantially increased since 2011 (Figure S5).

### Suicide trends by method

Table 1 summarizes suicide mortality rate trends by method, age group, and sex for the 2020-2024 period.

**Table 1:** Average annual percent change trends in suicide mortality rates by method, age group, and sex, 2020-2024.

| Method | Overall | 10-14 years |  |  | 15-19 years |  |  | 20-24 years |  |  |
| --- | --- | --- | --- | --- | --- | --- | --- | --- | --- | --- |
|  |  | All | Female | Male | All | Female | Male | All | Female | Male |
| **All** | ↓*** | ↓* | ↑* | ↓** | ↓** | ↓ | ↓*** | ↓* | ↓ | ↓** |
| **Firearms** | ↓ | ↑ | ↑** | ↓ | ↓* | ↑ | ↓ | ↑ | ↑** | ↓ |
| **Asphyxiation** | ↓*** | ↓* | ↑ | ↓*** | ↓*** | ↓** | ↓*** | ↓*** | ↓** | ↓*** |
| **Poisoning** | ↑ | ↑ | ↑ | NA | ↑*** | ↑** | ↑** | ↑ | ↑** | ↑ |
| **Other** | ↑*** | NA | NA | NA | ↑ | ↓ | ↑** | ↑*** | ↑*** | ↑*** |

Arrows indicate the directionality of the suicide mortality rate per 100,000 people, for each age and sex subgroup. Trend: ↑ = increasing; ↓ = decreasing. Trend estimates were based on the AAPC for 2020-2024. NA=Not available because the NCHS does not report suicide mortality rates for subgroups with death counts <10 to protect personal privacy and prevent identification. . \* for $p < 0.05$ , \*\* for $p < 0.01$ , \*\*\* $p < 0.001$ .

Since 1999, firearms have consistently accounted for the largest proportion of adolescent suicide deaths across all methods (54.2% in 2024; Figure 3 and Figure S2 for suicide death proportions over time). Firearm suicide rates increased from 3.2 to 5.9 per 100,000 from 2008 to 2021—a 84% rise and the largest increase of any suicide method over this period, peaking in 2021. From 2020-2024, firearm suicide rates did not significantly change (AAPC= −0.6 (CI: −3.2 to 1.9, p= 0.559). In 2024, the crude rate owing to firearms was 5.3 (CI: 5.1-5.5).

**Figure 3:**
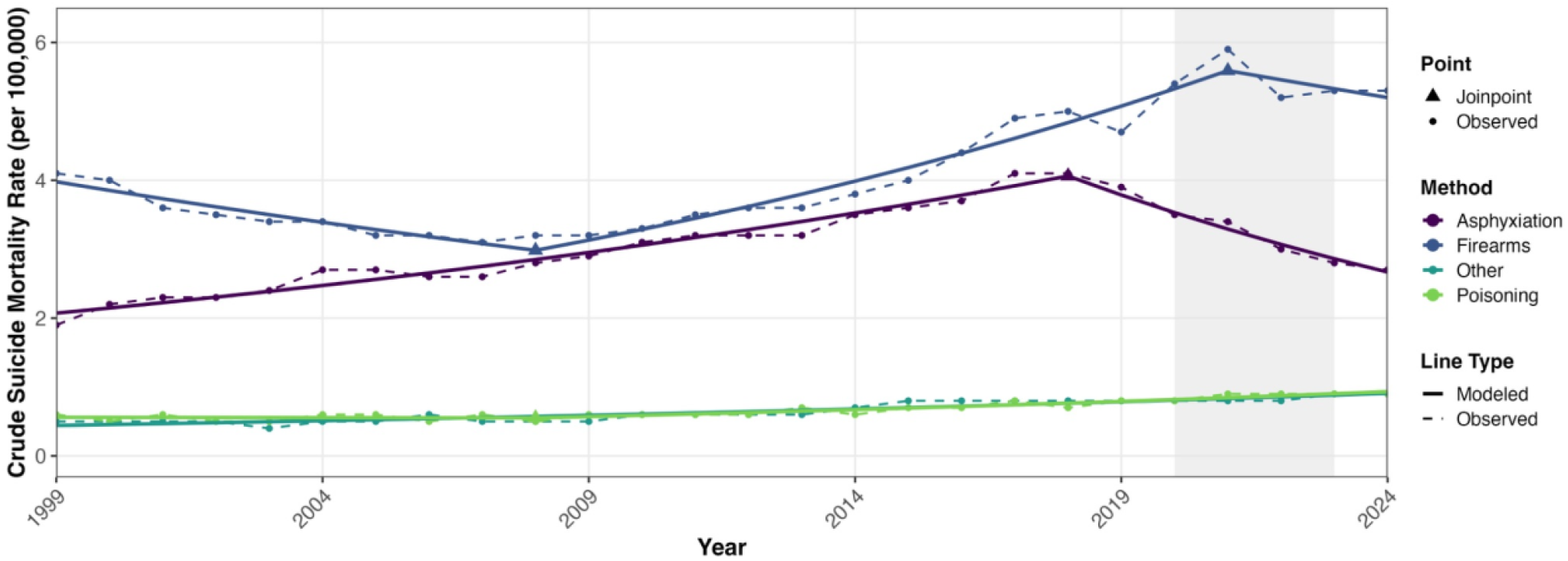
Joinpoint Regression of Crude Suicide Mortality Rates by Method for 10-24-year-olds. The grey shading denotes the COVID-19 emergency period (2020-2023) for the US. For the analysis restricted to adolescents aged 10-19, see Figure S3.

Meanwhile, asphyxiation was the second most common suicide method from 1999-2024 (CR 2024: 2.7 (2.6-2.8), 27.8% of all adolescent suicide deaths), with a peak in 2018. However, from 2020 to 2024, asphyxiation suicide rates decreased on average by 6.8 per year (CI: −8.5 to −5.0, p<0.001).

Finally, trends among less common methods indicate disparate trajectories. Though suicide rates by poisoning (CR 2024: 0.9 (0.8-1.0), 9.0% of all adolescent suicide deaths) did not change significantly (AAPC= 3.3, CI: −1.3 to 8.3, p=0.055), suicide rates by other means (CR 2024: 0.9 (0.8-0.9), 9.0% of suicide deaths) showed a steady rise over the past two decades (AAPC 2020-2024= 2.9 (CI: 2.3 to 3.6, p<0.001)).

#### Death by firearms, stratified by age and sex

Since 2015, firearms have been the most common method among individuals aged 15-19 and 20-24, accounting for 50.5% and 59.0% of suicides in these age groups, respectively, in 2024 (Figure 3). In the 10-14-year age group, despite asphyxiation-related deaths being more common, firearms still accounted for 37.8% of suicide deaths. Whereas firearm suicide mortality rates decreased by 1.3 on average since 2020 for ages 15-19 per year (CI: −3.1 to −0.0, p=0.046), they did not change significantly for ages 10-14 (AAPC 2020-2024= 2.0 (CI: −1.3 to 4.4), p=0.202) and ages 20-24 (AAPC 2020-2024= 0.1 (CI: −2.0 to 1.8), p=0.929).

Firearm use has consistently been more prevalent among males (61.9% of male suicides in 2024) than among females (29% of female suicides). The steep increase in firearm suicide mortality rates was mainly driven by suicides in males, among whom rates rose sharply until 2021(APC 2013-2021: 5.9 (CI: 4.9 to 9.1, p<0.001) and then decreased by 3.6 annually since 2021 (CI: −8.5 to −0.4, p=0.029).

In contrast, the rate among females sharply increased from 2007 to 2016 (APC 7.4 (CI: 5.6-16.4), p=0.01), after which it remained stable (APC 2016-2024: 2.5 (CI: −1.4 to 4.2), p=0.109). Recent increases in female firearm suicide rates are mainly attributable to ages 10-14 and 20-24 (Figure S6).

#### Death by asphyxiation, stratified by age and sex

From 2020 to 2024, the asphyxiation suicide mortality rates decreased for all age groups and for both sexes (Figure 4, 5, S6). However, heterogeneity by sex and age exists: asphyxiation has consistently been the predominant suicide method among females since 2001 (40.0% of suicides in 2024) and in the 10-14 age group across sexes since 1999 (53.4% of suicides in 2024). Although the overall asphyxiation suicide rate decreased since 2020, rates in females (AAPC 2020-2024= −2.5 (CI: −4.9 to −0.8,p=0.014) and in the 10-14 age group (AAPC 2020-2024= −6.3 (CI: −12.6 to −1.4, p=0.017)) decreased less than in the other demographic groups. Among males, asphyxiation suicide rates decreased on average by −10.1 (CI: −13.3 to −7.4, p<0.001) from 2020 to 2024. Similarly, rates decreased by 7.4 in the 15-19 age group (AAPC= −7.4 (CI: −10.6 to −5.0, p<0.001)) and by 8.9 in the 20-24 age group (AAPC= −8.9 (CI: −11.2 to −6.8, p<0.001)) across sexes.

**Figure 4:**
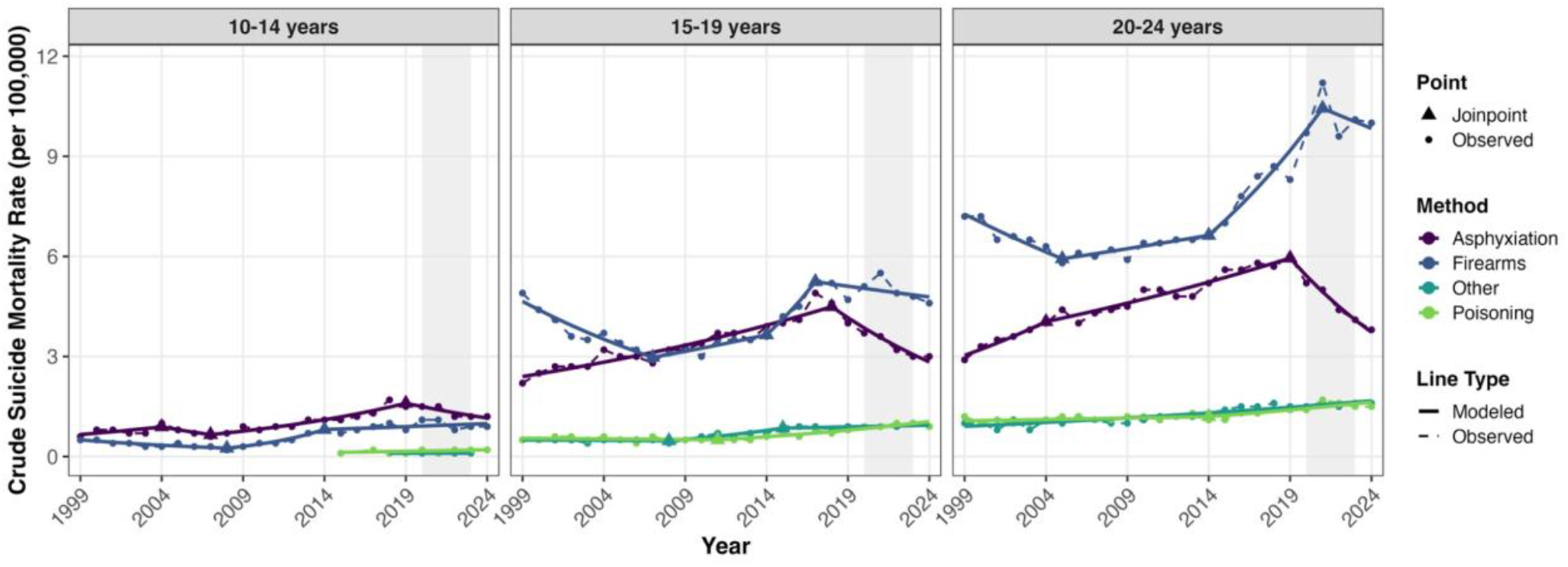
Trends in Crude Suicide Mortality Rates among Adolescents, by Method and Age Group. The grey shading denotes the COVID-19 emergency period for the US (2020-2023). Of note, in the 10-14 age group, data for several years were suppressed for suicides by poisoning and by other means (poisoning: 1999-2014, 2016; other: <2018, 2024) since counts were <10.

**Figure 5:**
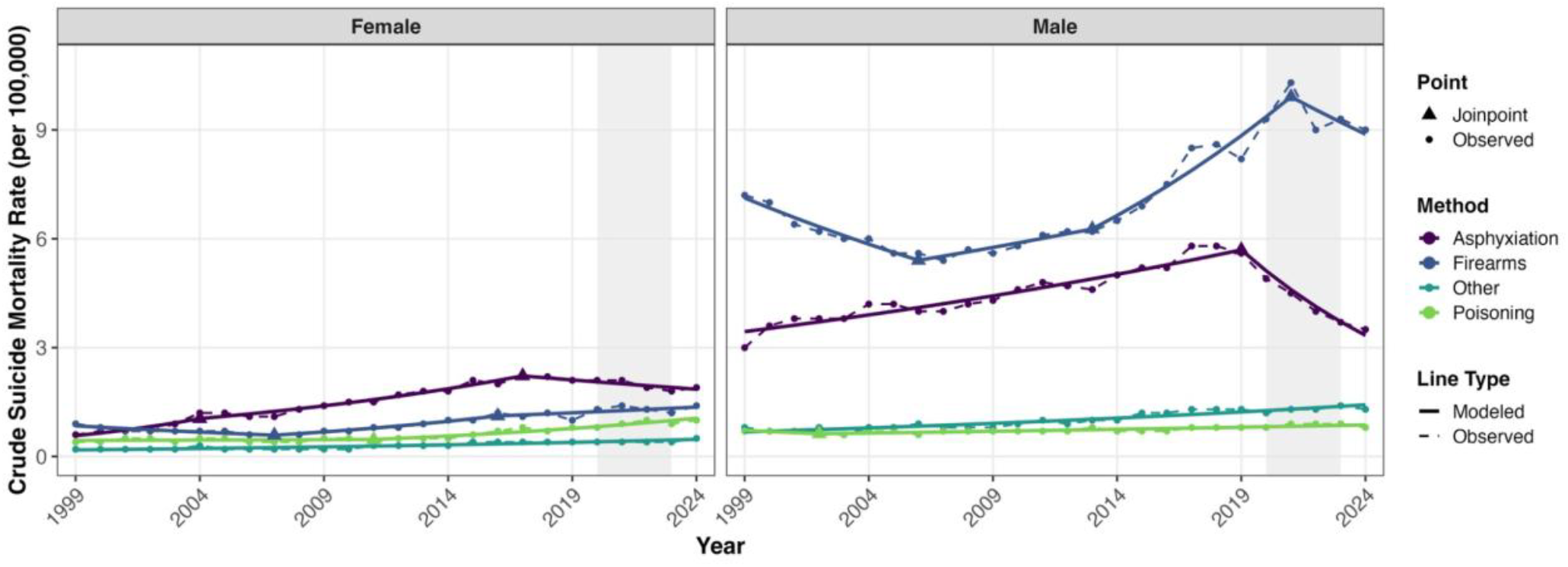
Trends in Crude Suicide Mortality Rates among Adolescents, by Method and Sex. The grey shading denotes the COVID-19 emergency period (2020-2023) for the US.

#### Death by poisoning, stratified by age and sex

Among individuals aged 15-19 and 20-24, poisoning suicide rates steadily increased since 2015, with an especially pronounced increase in the 15-19 age group (AAPC 2020-2024: 5.8 (CI:4-1 to 10.6, p<0.001). In 2024, suicides by poisoning accounted for 7.8% of suicide deaths in the 10-14 age group, 9.8% in the 15-19 age group, and 8.8% in the 20-24 age group.

While there was a pronounced increase among females (AAPC 2020-2024= 6.4 (CI: 4.3 to 14.1), p=0.018), poisoning suicide rates among males did not change significantly (AAPC 2020-2024= 1.5 (CI: −2.3 to 4.4), p=0.075) (Figure 5). As a result, in 2024, poisoning suicide deaths accounted for 20.9% of female deaths and 7.4% of male deaths. Deaths among individuals aged 10-14 were suppressed for all years except 2015 and 2017-2024 owing to small counts <10.

#### Death by other means, stratified by age and sex

Suicide mortality rates for other means – including jumping – have been increasing since 1999, with a slightly more pronounced rise in females (AAPC 2020-2024= 3.9 (CI: 3.1 to 4.9), p<0.001) than in males (AAPC 2020-2024= 3.0 (CI: 2.5 to 3.7), p<0.001). In 2024, death by other means comprised 9.7% of female and 12.1% of male suicide deaths. Stratifying by age group, other suicide mortality rates remained stable in the 15-19 age group (AAPC= 1.1 (CI: −1.2 to 2.5), p=0.268), while they increased by 2.5 ((CI: 1.9 to 3.1), p<0.001) in the 20-24 age group from 2020-2024. Deaths among individuals aged 10-14 were suppressed for all years except 2018-2023 owing to small cell counts.

Our results were robust to the use of age-standardized suicide mortality rates rather than crude rates.

## Discussion

Suicide mortality rates declined from 2020 to 2024 across age groups and for both sexes. This overall decrease was driven primarily by falling asphyxiation across sexes and by declining firearm suicide rates among males. Female suicide rates, however, did not follow this downward trend. Of particular concern is the rise in suicide rates among females in the age group 10-14, especially driven by firearm use. Rising rates of suicide by other means and poisoning in the age group 15-19 also warrant close attention.

Similar to adult firearm suicide trends, adolescent firearm suicide mortality rates rose dramatically during the COVID-19 emergency period and decreased again in more recent years.^15,16^ Still, firearms have remained the predominant suicide method among male adolescents throughout the entire 1999-2024 study period. Relatedly, trends among females have shown a gradual shift toward more lethal methods (i.e., decreasing share of deaths by asphyxiation alongside an increasing share of deaths by firearms)—a pattern consistent with findings reported through 2020.^4,16^ With prior research showing that most adolescents dying by firearm suicide used a gun belonging to a family member, secure storage, extreme risk protection orders, and youth access restriction remain a priority.^17,18^

Departing from prior observations, asphyxiation-related mortality rates have declined since 2019 across age groups and sexes. This may be partly explained by reduced access to isolated settings outside the home during the COVID-19 emergency period, where stay-at home orders, school closures, and potential increased supervision by family members may have limited opportunities for asphyxiation in private and unsupervised spaces Corroborating previous studies^4,19^, the continued increase in poisoning suicide rates in ages 15-19 and 20-24 is highly concerning. This may reflect facilitated access to poisons and lethal dosing information through online sources, including increased opportunities of ordering drugs online since COVID-19. Similarly, less conventional methods, including jumping, have steadily increased across both sexes and the 15–24 age group, in line with previously observed trends.^4^

Despite decades of research on the effectiveness of suicide prevention programs, the evidence base, particularly for adolescents, remains inconsistent. Across multiple systematic reviews and meta-analyses, structural interventions—especially means restriction—have shown the most robust effects, while school- and community-based programs have mainly demonstrated improvements in knowledge and attitudes rather than suicidal behavioral outcomes.^20–26^ Importantly, most of this evidence predates the COVID-19 pandemic and the rapid expansion of social media and generative artificial intelligence (AI)—contexts that may have fundamentally reshaped adolescent suicide risk. Recent reports about cases in which adolescent interactions with AI chatbots were alleged to have contributed to suicides,^27,28^ highlight how emerging technologies may introduce previously unknown risks. Viral challenges and algorithmically amplified content on social media platforms can blur the line between risk-taking behavior and suicidal intent.^29,30^ This shows that suicide risk programs need to be adapted accordingly.

In line with a meta-analysis suggesting that existing programs are more effective for men,^25^ our analyses highlight that prevention efforts to date have mainly shown effectiveness among male adolescents. Mann et al. concluded in 2021 that further suicide reduction requires evaluating newer approaches,^23^ including internet-based screening. Our findings confirm this urgent need and show that priority must now shift to developing and rigorously evaluating strategies tailored to female adolescents, particularly in the 10-14 age group.

### Limitations

With underresourced local death investigation systems^31^ and the ongoing polysubstance use epidemic,^32,33^ our investigation may be affected by suicide underreporting or misclassification of death certificates (e.g., accidental poisoning rather than intentional self-harm), especially in coroner-only states or in cases of unclear suicide intent.^34^ Our analysis did not stratify by race or ethnicity because further stratification resulted in substantial data suppression, although examining potential racial disparities will be a valuable step for future studies.

### Conclusion

Overall, suicide mortality rates declined from 2020 to 2024 among individuals aged 10-24 years, mainly driven by a reduction in asphyxiation use and a decrease in male suicide rates. Concerningly, female suicide rates did not follow this trend, with rising rates observed in individuals aged 10-14. Additionally, increasing suicide rates by other means and poisoning in specific subgroups warrant close attention. These trends highlight the need to revisit existing prevention strategies. Efforts to date appear to have been effective for males across methods and reductions in asphyxiation use across sexes. Priority should now shift toward female-focused prevention strategies, particularly for 10-14 year-olds. Refocusing suicide prevention efforts through targeted, sex- and age-sensitive approaches will be key to addressing the diverging trends observed across demographic groups.

## Data Availability

All data used in these analyses are openly available on the CDC WONDER platform. All supplementary materials and analysis code are available upon reasonable request to the authors.

https://wonder.cdc.gov/

## Authors’ contributions

The authors contributed to the study in the following ways: conceptualization and study design were carried out by ACB, MC, KDM, and MSM; data collection, analysis, and visualization were conducted by ACB; validation was performed by ACB, MC, KDM, and MSM; the initial manuscript was drafted by ACB; and the manuscript underwent critical review and revisions by MC, KDM, and MSM. All authors had complete access to all study data, approved the final manuscript, and accepted responsibility for its submission for publication.

## Funding

This work was supported by the National Science Foundation (IIS2229881) and the National Institutes of Health (R35GM146974), as well as the German National Academy of Sciences Leopoldina through a postdoctoral fellowship grant (LPDS 2024-06) awarded to Anne Bischops. In partial support of M-LC work, MSM’s lab has recently been awarded funds for a postdoctoral fellowship from Moderna. The funders had no role in study design, data collection, data analysis, data interpretation, or writing of this work.

## Declaration of interests

The authors declare no competing interests.

## Notes

### Competing Interest Statement

The authors have declared no competing interest.

### Author Declarations

CDC WONDER platform

